# Hydroxychloroquine for pre-exposure prophylaxis of COVID-19 in health care workers: a randomized, multicenter, placebo-controlled trial (HERO-HCQ)

**DOI:** 10.1101/2021.08.19.21262275

**Authors:** Susanna Naggie, Aaron Milstone, Mario Castro, Sean P. Collins, Lakshmi Seetha, Deverick J. Anderson, Lizbeth Cahuayme-Zuniga, Kisha Batey Turner, Lauren W. Cohen, Elizabeth Fraulo, Anne Friedland, Jyotsna Garg, Anoop George, Hillary Mulder, Rachel E. Olson, Emily C. O’Brien, Russell L. Rothman, Elizabeth Shenkman, Jack Shostak, Christopher W. Woods, Kevin J. Anstrom, Adrian F. Hernandez, on behalf of the HERO Research Program

## Abstract

**Objective:** To determine whether hydroxychloroquine (HCQ) is safe and effective at preventing COVID-19 infections among health care workers (HCW).

**Design:** Multicenter, 1:1 randomized, placebo-controlled, double-blind, parallel-group, superiority trial.

**Setting:** 34 clinical centers in the United States.

**Participants:** 1360 HCW at risk for COVID-19 infection enrolled between April and November 2020.

**Interventions:** A loading dose of HCQ 600 mg twice on Day 1 followed by 400 mg daily for 29 days or matching placebo taken orally.

**Main Outcome Measure:** Composite of confirmed or suspected COVID-19 clinical infection by Day 30 defined as new onset fever, cough, or dyspnea and either a positive SARS-CoV-2 PCR test (confirmed) or a lack of confirmatory testing due to local restrictions (suspected).

**Results:** Enrollment for the study was closed before full accrual due to difficulties recruiting additional participants. The primary composite endpoint occurred in 41 (6.0%) participants receiving HCQ and 53 (7.8%) participants receiving placebo. No statistically significant difference in the proportion of participants experiencing clinical infection (estimated difference of -1.8%, 95% confidence interval -4.6% to 0.9%, p=0.20). We identified no significant safety issues.

**Conclusion:** Oral HCQ taken as prescribed appeared to be safe in a group of HCW. No significant clinical benefits were observed. The study was underpowered to rule out a small but potentially important reduction in COVID-19 infections.

**Trial Registration:** NCT04334148

## Introduction

After the emergence of SARS-CoV-2 in late 2019, the virus spread rapidly, resulting in the worst pandemic in nearly a century. Early in the pandemic, health care systems struggled with maintaining adequate supply of personal protective equipment (PPE), and infections in health care workers (HCW) were reported globally.^1^ Without availability of protective vaccines, there was a need to identify therapies that might prevent infection and could be taken regularly by HCW who were at high risk of frequent exposures, similar to approaches taken with malaria and human immunodeficiency virus pre-exposure prophylaxis. As is common with new diseases, repurposed drugs offered immediate options for therapeutics and usually with a well-known safety profile, allowing for more rapid introduction into clinical trials and thereafter into clinical practice.

Chloroquine had been previously reported to have *in vitro* antiviral activity against SARS-CoV-1 and MERS-CoV, and both chloroquine and hydroxychloroquine (HCQ) showed similar *in vitro* activity against SARS-CoV-2.^2-4^ Thus, clinicians turned to HCQ early in the pandemic as a therapy that might have clinical benefit for treating COVID-19, the disease associated with SARS-CoV-2 infection, as investigators began testing the drug’s safety and efficacy in the treatment and prevention of COVID-19. While vaccines are now available for the prevention of SARS-CoV-2 infection, access to sufficient quantities of vaccine remains challenging, particularly in low- and middle-income countries. Vaccine hesitancy further complicates the public health response. Thus, preventative therapy for SARS-CoV-2 infection remains relevant.

The Healthcare Worker Exposure Response and Outcomes (HERO) Registry (NCT04342806) was created as a community of HCW from across the United States to learn about issues impacting frontline workers and to offer opportunities to participate in research studies.^5^ The HERO-HCQ trial was one of the first studies in the United States to test the safety and efficacy of HCQ as pre-exposure prophylaxis in frontline HCW. HERO-HCQ leveraged both the HERO Registry as well as its relationship with PCORnet®, The National Patient-Centered Clinical Research Network as a pragmatic and largely remote clinical trial, using a patient-facing online portal to capture frequent patient-reported outcomes.^6^ The primary objective of HERO-HCQ was to evaluate the efficacy of HCQ in preventing SARS-CoV-2 clinical infection in HCW when taken daily. Secondary objectives of the study were to assess the efficacy of HCQ in preventing asymptomatic viral shedding of SARS-CoV-2 among HCW and to assess the safety and tolerability of HCQ in this study population.

## METHODS

### Study Design

HERO-HCQ was a multicenter, double-blind, randomized, parallel-group study designed to evaluate the superiority of HCQ vs. placebo for COVID-19 pre-exposure prophylaxis in HCWs. The HERO-HCQ trial was reviewed by the Duke University School of Medicine Institutional Review Board and approved by the Western Institutional Review Board (Pro00105274).

### Participants

Eligible participants provided informed consent, were age 18 or older, and were working in a health care setting with potential exposure to patients with COVID-19. A detailed list of eligible health care settings is shown in the protocol **[Supplemental Table 1]**. The main exclusions were prior diagnosis of COVID-19 infection or contraindications to HCQ.

### Randomization and Masking

Participants were randomized in a 1:1 ratio to receive HCQ or placebo at the level of the individual participant via the study portal. Randomization was stratified by clinical site using a permuted block design with varying block sizes. In the intervention arm, participants received an oral loading dose of study drug at 600 mg twice on the first day, followed by 400 mg daily for 29 days. In the control arm, placebo tablets were administered using the same dosage schedule and number of tablets as the intervention arm. The placebo was similar in appearance to the study drug and packaged and labeled in a masked manner in compliance with regulatory requirements. All study drug doses were oral self-administrations. Study drug was supplied as 200-mg tablets, and each eligible participant was provided a quantity sufficient for 30 days.

### Procedures

Participants were prescreened through the HERO Registry, and eligibility was confirmed by the site by phone or in person.^5^ Participants were able to electronically consent through the portal, which was done at the time of the site visit or in advance. There were two on-site visits – one at baseline and another at 30 days. Baseline assessments included a nasopharyngeal swab for SARS-CoV-2 and a blood sample to assess baseline SARS-CoV-2 nucleocapsid IgG antibody status. Weekly follow-up was performed remotely via standardized questionnaires utilizing the online portal. These questionnaires included screening for COVID-19 clinical signs/symptoms and self-reporting of COVID-19 testing and diagnosis. Additionally, participants were able to self-report medication changes, hospitalizations, clinical events, and adverse events. A call center provided support for missed visits to re-engage and remind participants to complete the questionnaires.

The second on-site visit at approximately 30 days was completed to assess study drug adherence and any subsequent clinical or safety events. A nasopharyngeal swab for SARS-CoV-2 PCR and a blood sample for SARS-CoV-2 nucleocapsid IgG antibody were obtained. Individual participants received study drug for 30 days and were followed for an additional 30 days for clinical events and patient-reported outcomes. An end-of-study virtual visit was conducted approximately 60 days after randomization via the direct-to-participant portal or call center to assess for any subsequent clinical or safety events.

### Outcomes

The primary outcome was a composite of confirmed or suspected clinical infection with COVID-19 through 30 days, which was defined as new onset of fever, cough, or dyspnea and confirmed SARS-CoV-2 PCR positive test result via local testing or suspected COVID-19 disease without confirmation testing due to local restrictions or policies. Participants who developed symptoms of COVID-19 were expected to follow local clinical and/or employee health protocols for testing and management. Secondary outcomes were (1) viral shedding of SARS-CoV-2 at 30 days and (2) safety and tolerability as determined by subject-reported adverse events that met criteria per protocol for serious adverse events and HCQ-associated events of special interest; this latter group comprised fever, arrhythmia (ventricular), psychosis, angioedema, prolonged QT interval, secondary bacterial infection, and suicidal ideation.

Exploratory outcomes were (1) SARS-CoV-2 nucleocapsid IgG seroconversion at 30 days; (2) COVID-19 complications including hospitalization, ICU level care, or need for invasive ventilation; (3) days sick or lost work time; (4) self-reported health and well-being obtained from the Patient-Reported Outcomes Measurement Information System (PROMIS) Emotional Distress-Anxiety-Short Form^7^, a Single Item Burnout Measure^8-11^, and the Patient Health Questionnaire (PHQ-2)^12-13^; and (5) patient-reported clinical infections among household contacts and other impacts on HCW household.

### Statistical Analysis

The original sample size of approximately 15,000 randomized participants was selected to yield high power for testing the primary outcome of clinical infection with SARS-CoV-2 assuming the usual risk of SARS-CoV-2 infection is 5%. This sample size was expected to provide greater than 80% power to detect a 1% absolute decrease (20% relative decrease) in COVID-19 infection rates between treatment arms. These calculations assumed a two-sided Type I error rate of 0.05 with 1:1 randomization. In October 2020, due to slower than expected enrollment and changing community attitudes about HCQ effectiveness, the study protocol was amended to reduce the total sample size to 2000, which provided 80% power to detect a 50% relative decrease in the risk of COVID-19 infections assuming a placebo group risk of 5%.

Statistical comparisons were performed using two-sided significance tests. The primary endpoint was clinical infection with SARS-CoV-2 through the 30-day period. Data collected during the 60-day follow-up were included for the safety analyses. For the primary outcome of clinical infection with SARS-CoV-2, comparisons between treatment arms were presented as differences in proportions with 95% confidence intervals (CIs) using the Miettinen-Nurminen method and a p-value calculated using the Fisher’s exact test. A secondary analysis was based on a logistic regression model with an indicator variable for the treatment group. Supplemental analyses were conducted to (1) examine the differences using other methods for constructing the Cis^14^ and (2) compute the common odds ratio using the Mantel–Haenszel test stratifying by enrolling site.^15^

Subgroup analyses were planned for age, sex, race and ethnicity. For each subgroup analysis, a logistic regression model was estimated, with additional terms identifying subgroup membership and intervention by subgroup interaction. The statistical comparisons of serious adverse events and events of special interest were based on chi-square tests and Fisher’s exact test. All analyses were conducted using SAS Version 9.4 software. The DCRI served as the Statistical and Data Coordinating Center.

### Patient and Public Involvement

HCW were engaged in the HERO program and trial through membership in HERO governance, including participation in the steering committee and subcommittees. HCW stakeholders reviewed enrollment materials and the study protocol, and advised on trial conduct throughout the study. An independent data safety monitoring board, which included an HCW representative, met regularly and monitored participant safety and study performance. The protocol as publicly shared and is available on heroesresearch.org.

## RESULTS

The HERO-HCQ trial start-up timeline was 4 weeks from concept to first participant randomized **(Supplemental Figure 1)**. Between April 2020 and November 2020, a total of 1360 participants were enrolled and randomized from 34 US sites participating in PCORnet **(Figure 1)**. One participant had a positive SARS-CoV-2 PCR test at the time of the baseline visit and was excluded from the primary analysis population. Overall, 92.9% and 92.3% of randomized participants completed their PCR and serology tests, respectively, at their Day 30 visit with no significant difference by treatment group. The Day 60 visit was completed by 89.0% of total participants.

**Figure 1.**
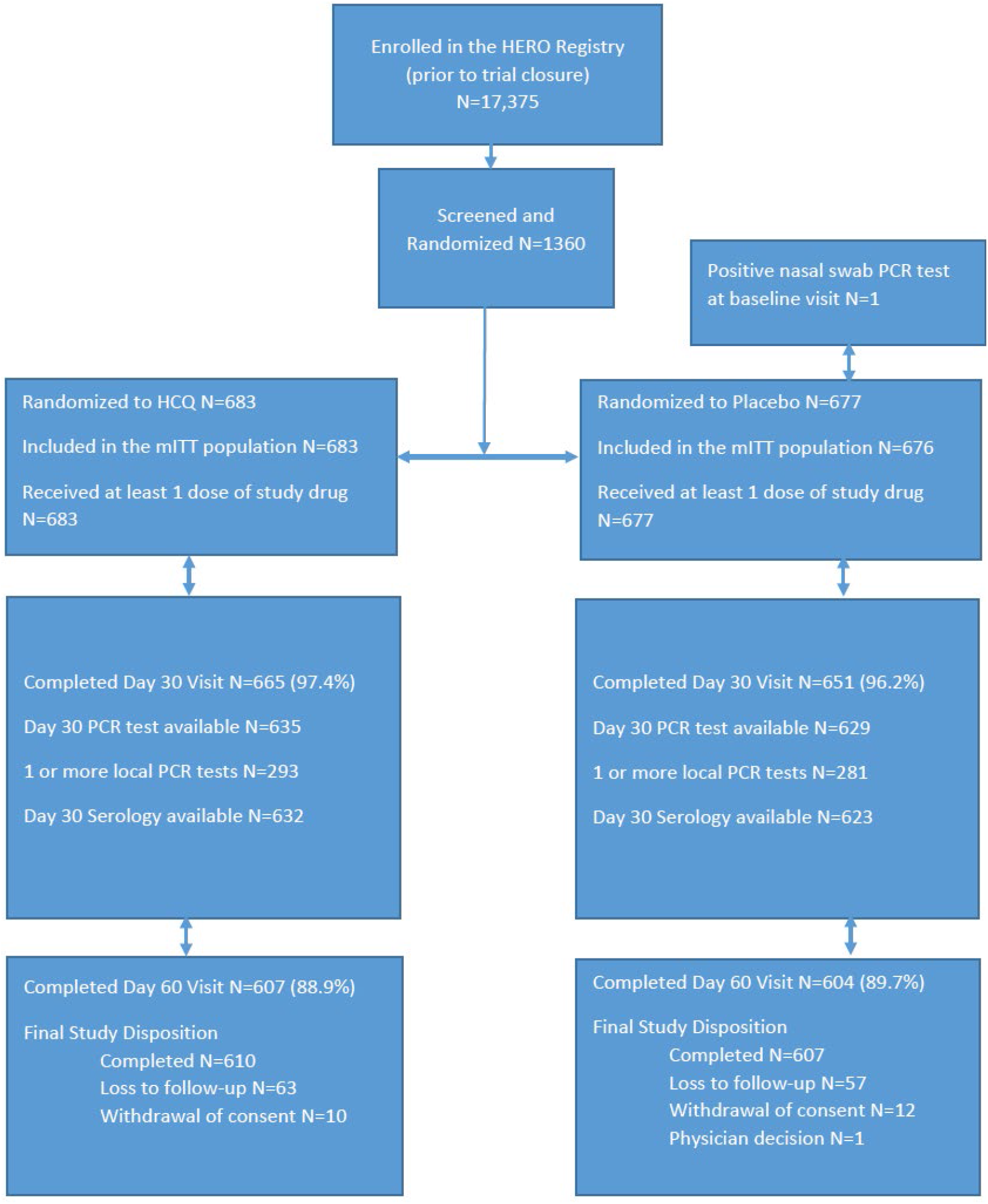
Enrollment, Randomization, and Follow-up.

### Baseline Participant Characteristics

The mean age of the HCW in the study population was 43.6 years, 65.3% were female, 90.8% reported being White race, and 5.8% self-reported as Hispanic or Latino ethnicity **(Table 1)**. The median body mass index was 27.1 kg/m^2^, and 33.2% of the population was considered obese (BMI ≥ 30 kg/m^2^). The most common comorbidities were hypertension (14.6%), asthma (9.9%), and diabetes (4.0%). Among the most common HCW locations were the emergency department (14.0%), ambulatory or outpatient care (9.5%), inpatient medical unit (8.5%), emergency medical services (8.1%), intensive care unit (7.9%), inpatient surgical unit (6.8%), and dedicated COVID-19 unit (5.7%). Among the enrolled participants, the most common occupation/employment characteristics were registered nurse (26.2%), physician (21.3%), nurse practitioner (5.2%), and paramedic (5.2%). Twelve (0.9%) participants were positive for SARS-CoV-2 nucleocapsid IgG at study enrollment.

**Table 1.** Baseline participant characteristics

| Characteristics | Hydroxychloroquine<br>(n=683) | Placebo (n=676) |
| --- | --- | --- |
| Mean age (SD) (years) | 44.2 (11.9) | 43.1 (11.2) |
| Women | 442 (64.7%) | 446 (66.0%) |
| Race or ethnicity |  |  |
| Black or African American | 18 (2.6%) | 23 (3.4%) |
| White | 624 (91.4%) | 610 (90.2%) |
| Other | 41 (6.0%) | 43 (6.4%) |
| Hispanic or Latino | 39 (5.7%) | 40 (5.9%) |
| Mean Weight (SD) (kg) | 82.5 (21.4) | 83.1 (21.8) |
| Median BMI (SD) (kg/m<sup>2</sup>) | 28.3 (6.3) | 28.6 (6.7) |
| Obesity | 226 (33.1%) | 225 (33.3%) |
| Hypertension | 99 (14.5%) | 99 (14.6%) |
| Asthma | 58 (8.5%) | 77 (11.4%) |
| Diabetes | 20 (2.9%) | 35 (5.2%) |
| COPD | 1 (0.1%) | 2 (0.3%) |
| Coronary artery disease | 5 (0.7%) | 6 (0.9%) |
| Occupation characteristics* |  |  |
| Registered nurse | 186/ 677 (27.5%) | 167/668 (25.0%) |
| Physician | 143/ 677 (21.1%) | 144/ 668 (21.6%) |
| Nurse practitioner | 33/ 677 (4.9%) | 37/ 668 (5.5%) |
| Paramedic | 30/ 677 (4.4%) | 40/ 668 (6.0%) |
| Qualifying hospital location* |  |  |
| Emergency department | 96 (14.1%) | 94 (13.9%) |
| Ambulatory care unit | 66 (9.7%) | 63 (9.3%) |
| Medical unit | 52 (7.6%) | 63 (9.3%) |
| Emergency medical services | 57 (8.3%) | 53 (7.9%) |
| Intensive care unit | 48 (7.0%) | 59 (8.7%) |
| Surgical unit | 50 (7.3%) | 43 (6.4%) |
| COVID-19 hospital unit | 38 (5.6%) | 39 (5.8%) |
| Positive SARS-CoV-2 nucleocapsid IgG<br>at study entry | 4/671 (0.6%) | 8/668 (1.2%) |
| Mean number of people living in the<br>home of the participant (SD) | 2.5 (1.4) | 2.5 (1.4) |

BMI = body mass index; COPD = chronic obstructive pulmonary disease.
\*Only characteristics with >5% are presented.

### Primary Endpoint

There were a total of 94 primary endpoint events during the 30-day follow-up period. Most of these endpoints were suspected clinical infection (n=85) rather than confirmed clinical infection with COVID-19 (n=9) **(Table 2)**. The most common presenting symptoms were cough (86.2%), fatigue (68.1%), headache (66.0%), and muscle aches/joint pain (51.1%). There were numerically fewer primary endpoint events in the HCQ group, 41 (6.0%), compared with the placebo group, 53 (7.8%); however, this difference of -1.8% (95% CI -4.6% to 0.9%) was not statistically significant (Fisher’s exact p=0.20) **(Supplemental Table 2)**. A secondary analysis based on a logistic regression model yielded similar results (OR 0.75, 95% CI 0.49 to 1.15, p=0.18). Among the participants with confirmed clinical infection, there were numerically fewer in the HCQ group (3 events [0.4%]) compared to the placebo group (6 events [0.9%]), and the difference was not statistically significant (0.45%, 95% CI -1.54% to 0.50**%**) (**Figure 2A**,**B)**. Supplemental analyses using the Mantel–Haenszel method, which stratified by enrolling site, yielded a similar estimate of the common OR (0.69, 95% CI 0.45 to 1.05). However, there was evidence of heterogeneity at the site level for the primary endpoint (p=0.011).

**Table 2.**
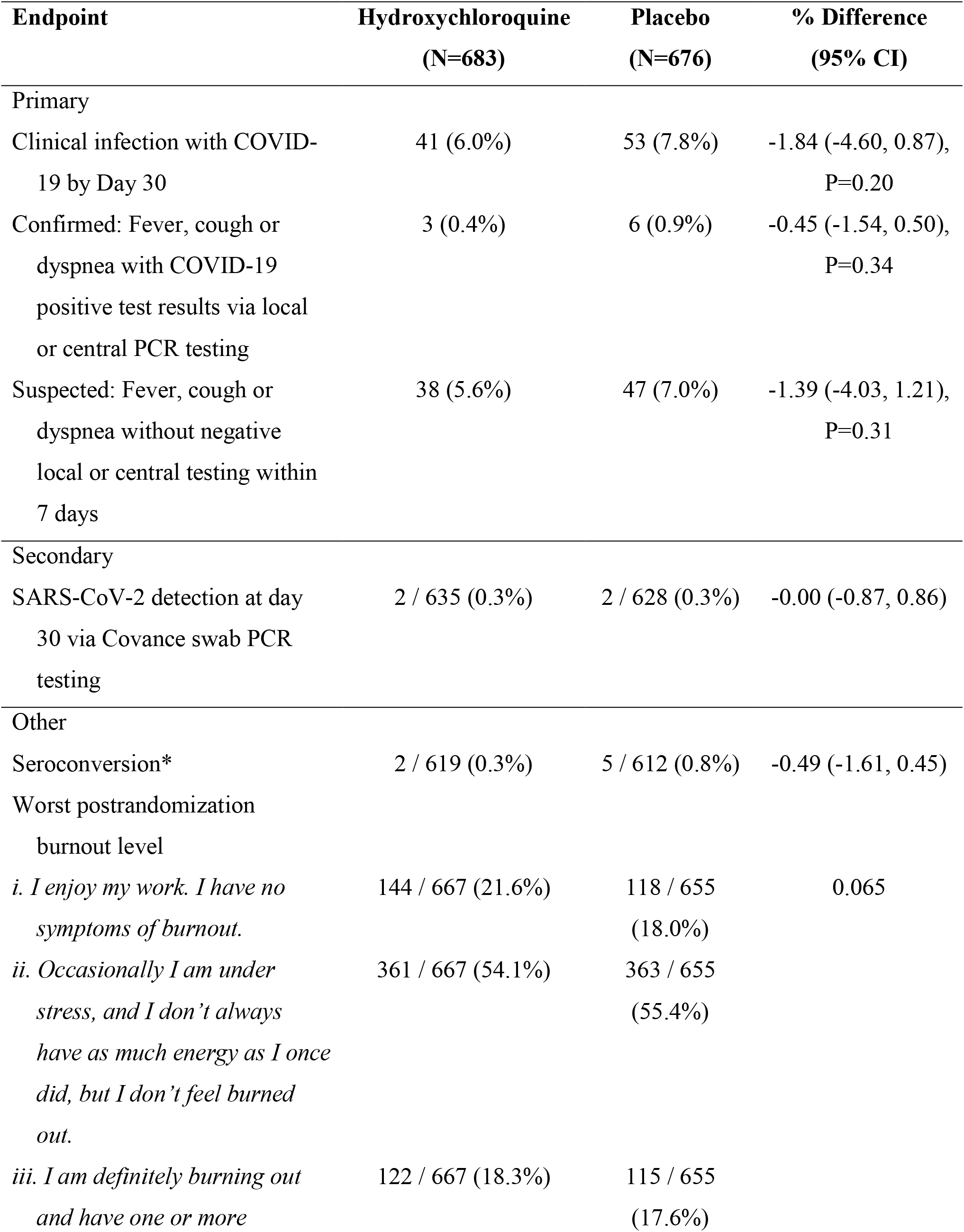
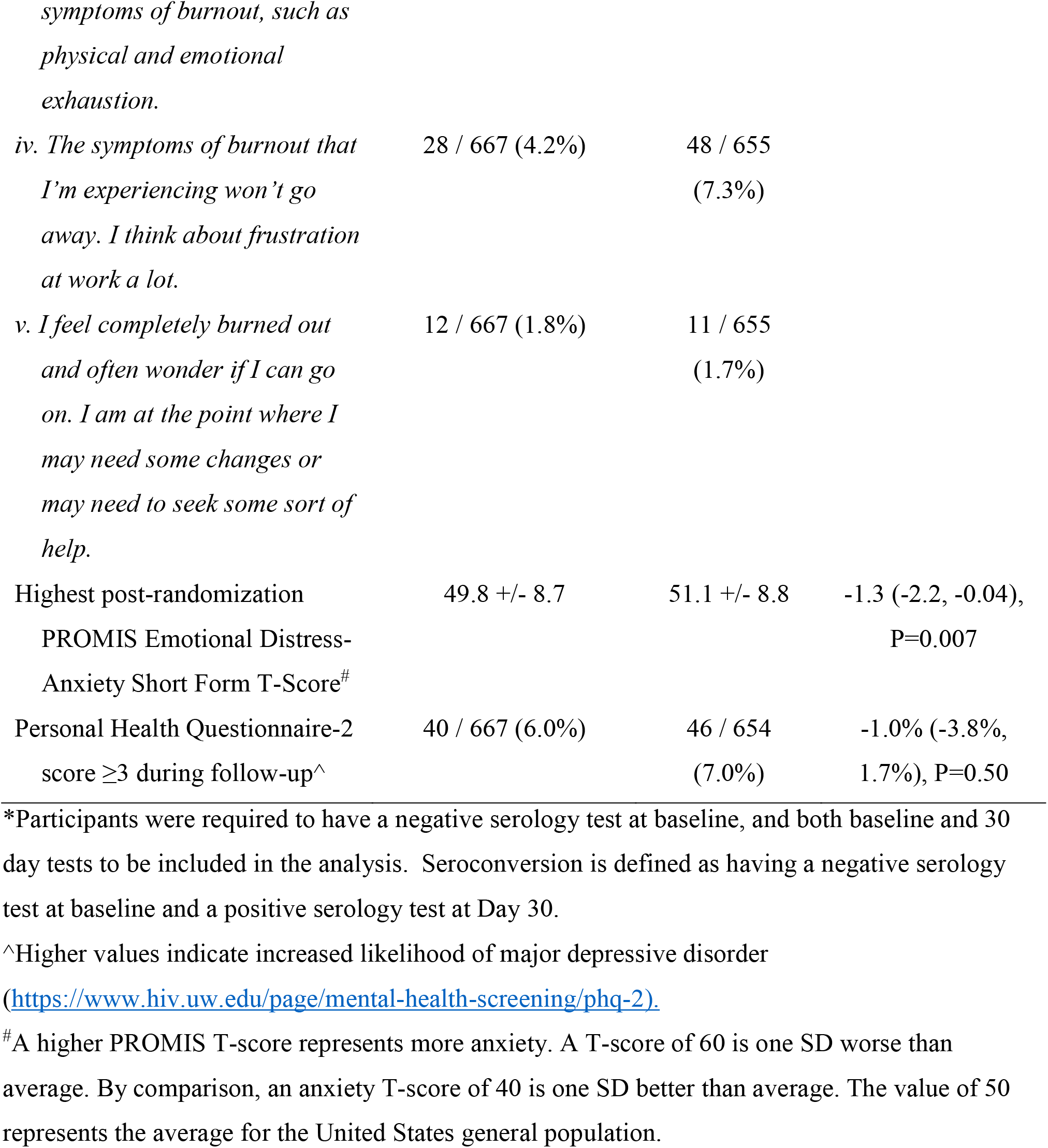
Key outcomes

**Figure 2.**
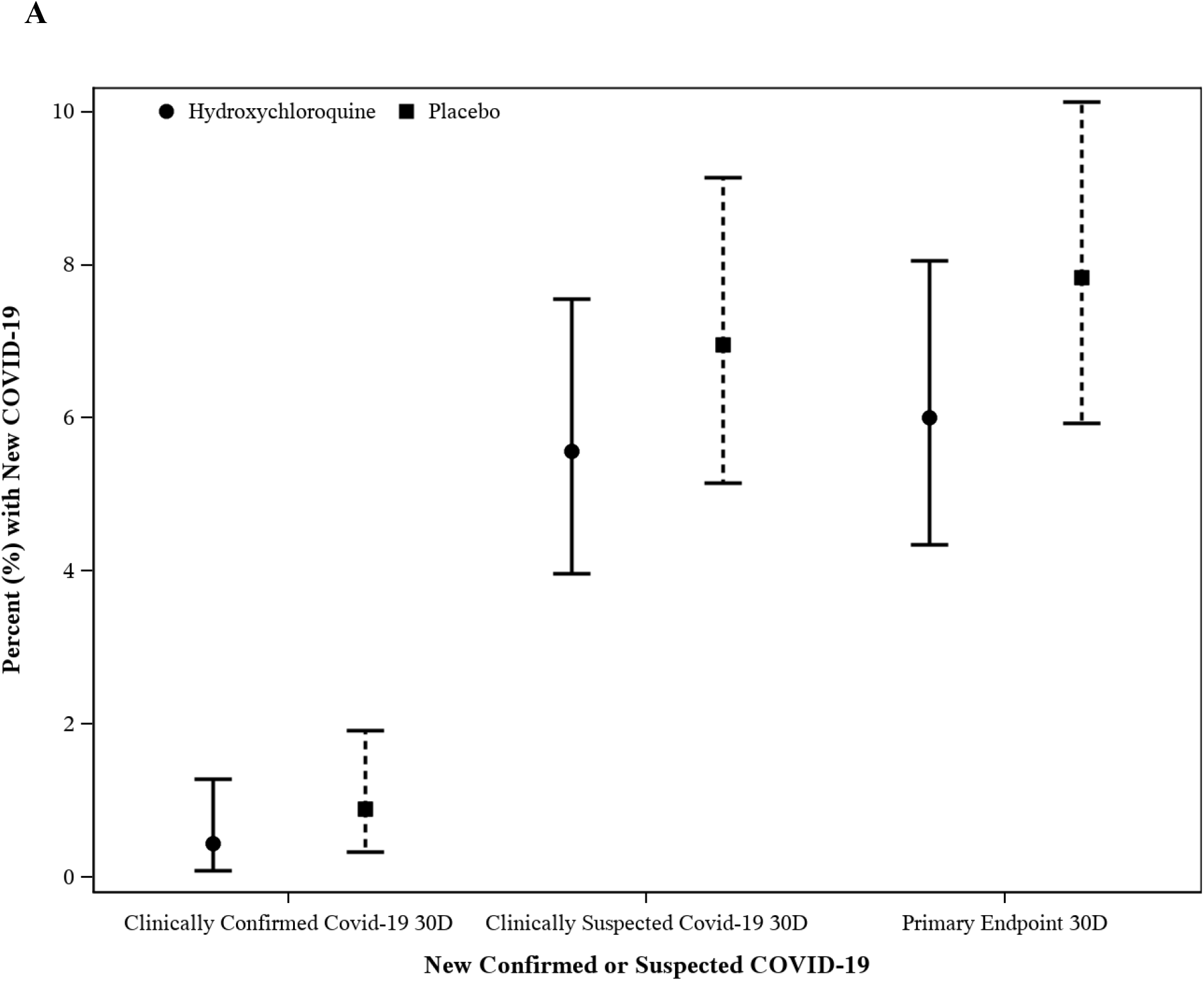
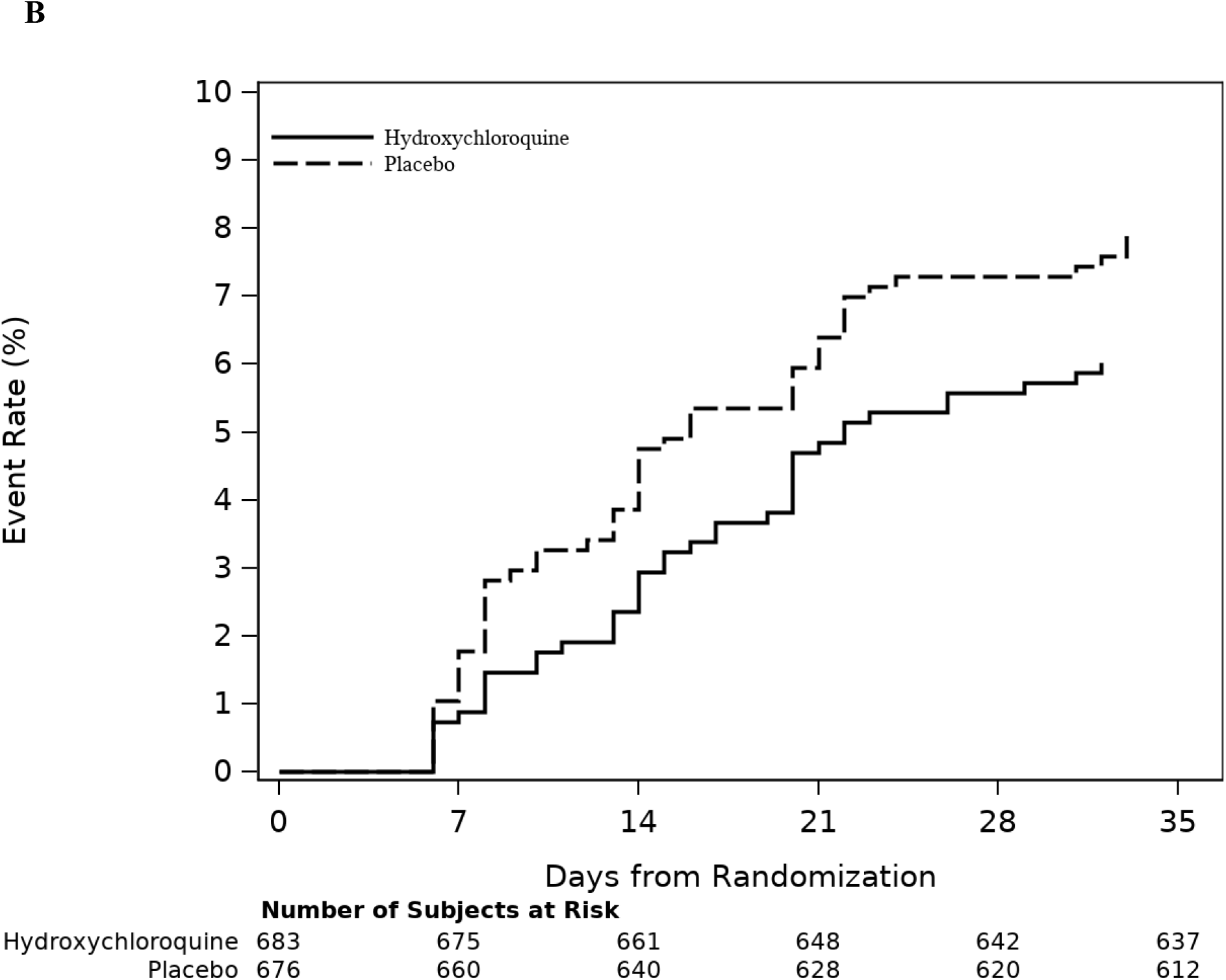
Primary Outcome, Overall and by Component. A, Confirmed, suspected, and overall primary endpoint by treatment group. B, Time-to-primary endpoint by treatment group.

### Subgroup Analyses

The prespecified subgroups for the primary endpoint are shown in **Figure 3**. All subgroups except for the youngest age group (18-35 years) showed estimated odds ratios <1.0 favoring HCQ. There were no subgroups suggesting heterogeneous treatment effect.

**Figure 3.**
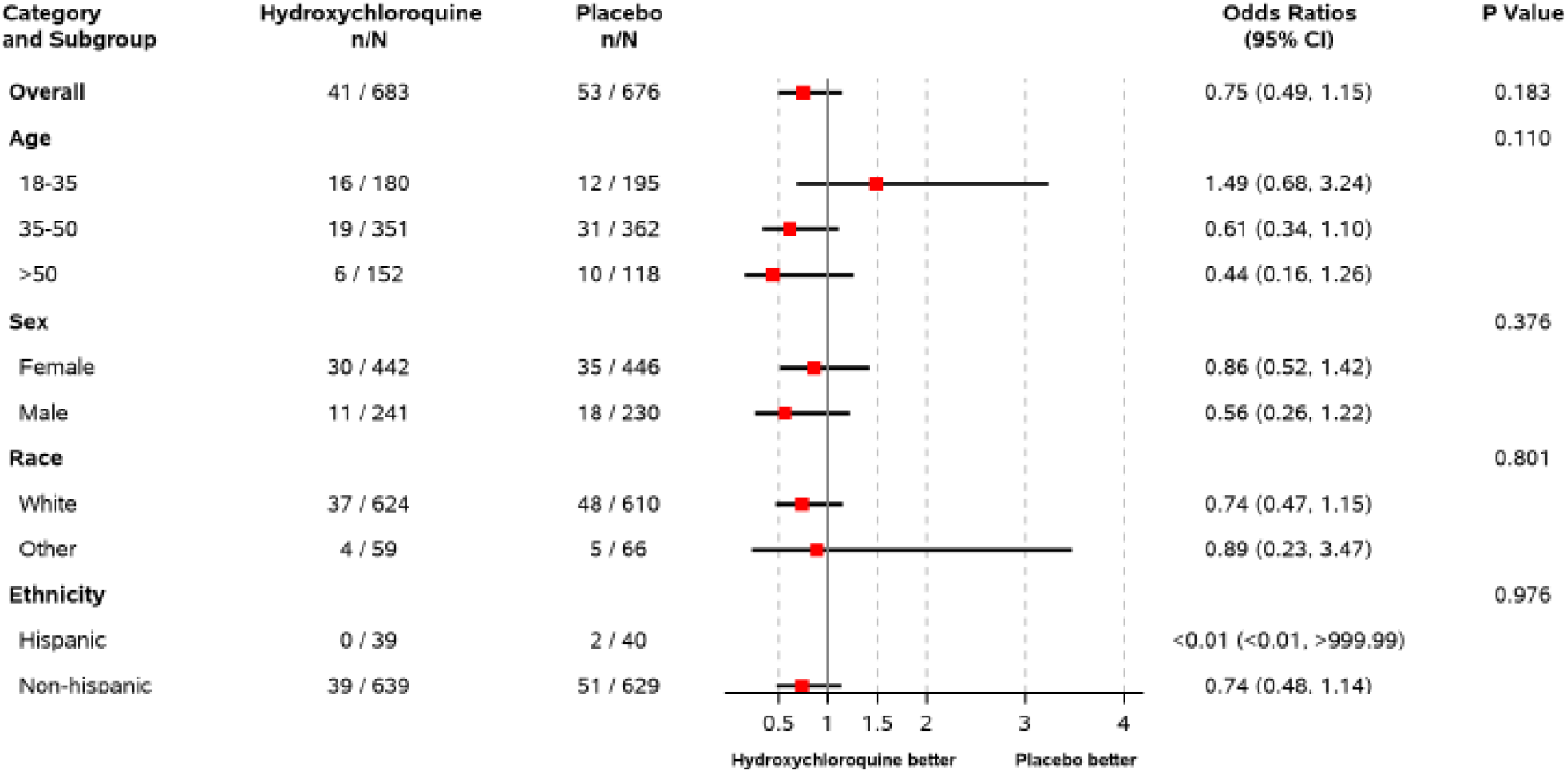
Prespecified Subgroup Analyses. Shown are the point estimates for the treatment effect on the odds ratio scale (95% CI).

### Secondary Endpoints

Four participants had a positive SARS-CoV-2 PCR test at the Day 30 visit, with each treatment arm having two positive cases (0.3% viral shedding rate at 30 days for both groups, p=1.00). Similarly, there were few seroconversions, defined as having a negative SARS-CoV-2 nucleocapsid IgG at entry and a positive IgG at Day 30, with two (0.3%) participants in the HCQ arm and five (0.8%) in the placebo arm having evidence of seroconversion. There were no deaths reported during the study period.

Participants in the HCQ arm reported statistically significantly lower levels of emotional distress and anxiety during the 30-day treatment period based on the worst recorded PROMIS Short Form T-scores (49.8 vs. 51.1 for a difference of -1.3 points, 95% CI -2.2 to -0.4, p=0.007). The percentage of participants reporting the PHQ-2 score ≥ 3 was not different between the two treatments (6.0% for HCQ vs. 7.0% for placebo; p=0.50). Levels of burnout were not statistically different between groups over the treatment period (p=0.065); however, numerically more participants in the HCQ arm reported no symptoms of burnout (21.6% vs. 18.0%) compared to placebo.

### Adherence to Treatment and Study Drug Discontinuation

All participants reported taking at least some study drug. At day 30, self-reported adherence (i.e., taking the drug for 29 days) was 94.4% for HCQ and 95.7% for placebo (p=0.32). Permanent discontinuation rates were 4.1% for HCQ and 2.7% for placebo. Permanent discontinuation due to adverse events was more common in the HCQ group (12 of 28 discontinuations) than the placebo group (3 of 18 discontinuations).

### Adverse Events and Safety

Adverse events of special interest (fever, ventricular arrhythmia, psychosis, angioedema, prolonged QT interval, secondary bacterial infection, and suicidal ideation) over the 60 days of follow-up were similar across groups (7 subjects in HCQ arm and 8 in placebo). At 60 days, subjects with serious adverse events were 3 (0.4%) for the HCQ group and 2 (0.3%) for the placebo group. Of those serious adverse events, 2 for the HCQ group and 1 for the placebo group resulted in hospitalizations **(Table 3)**.

**Table 3.**
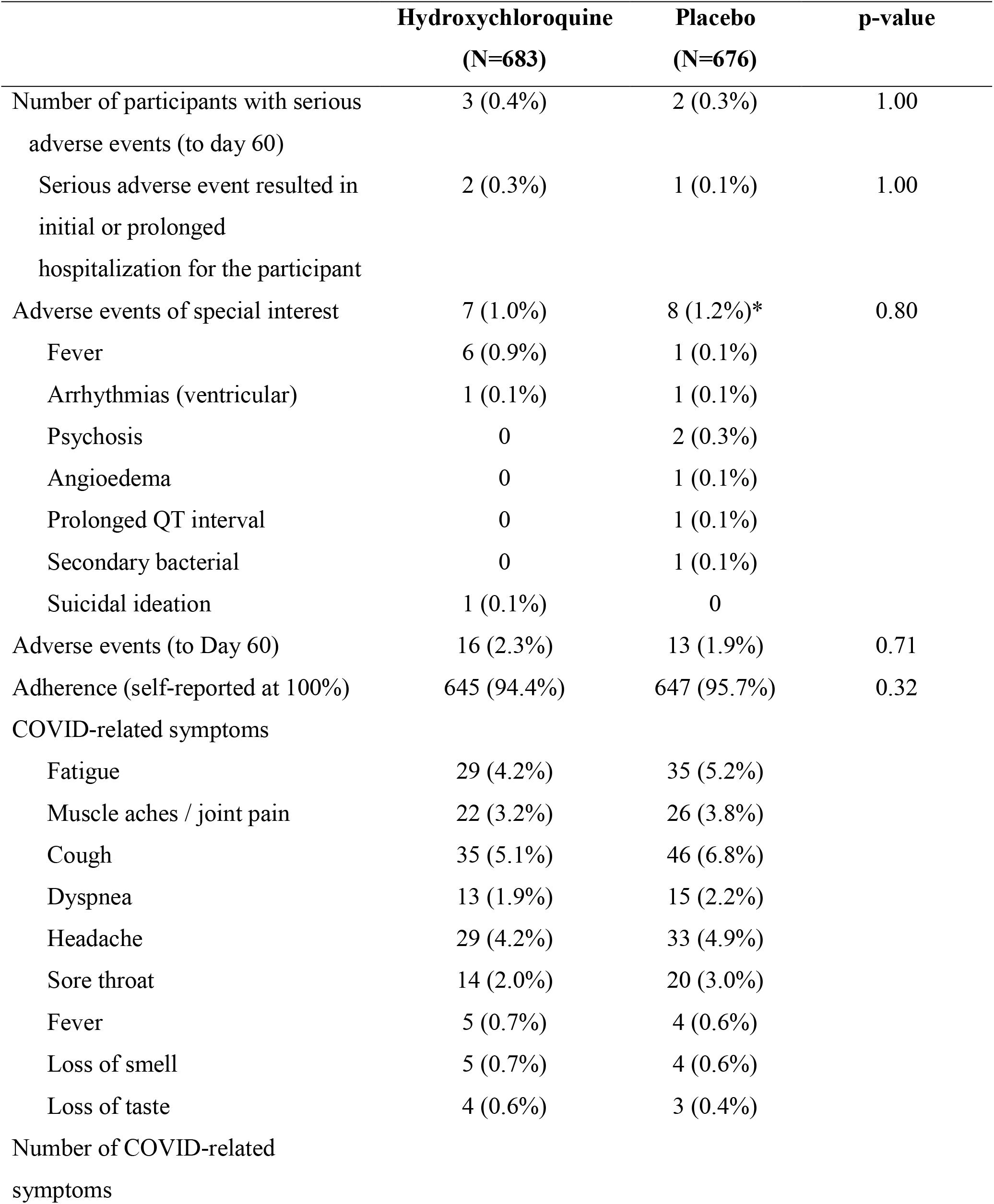
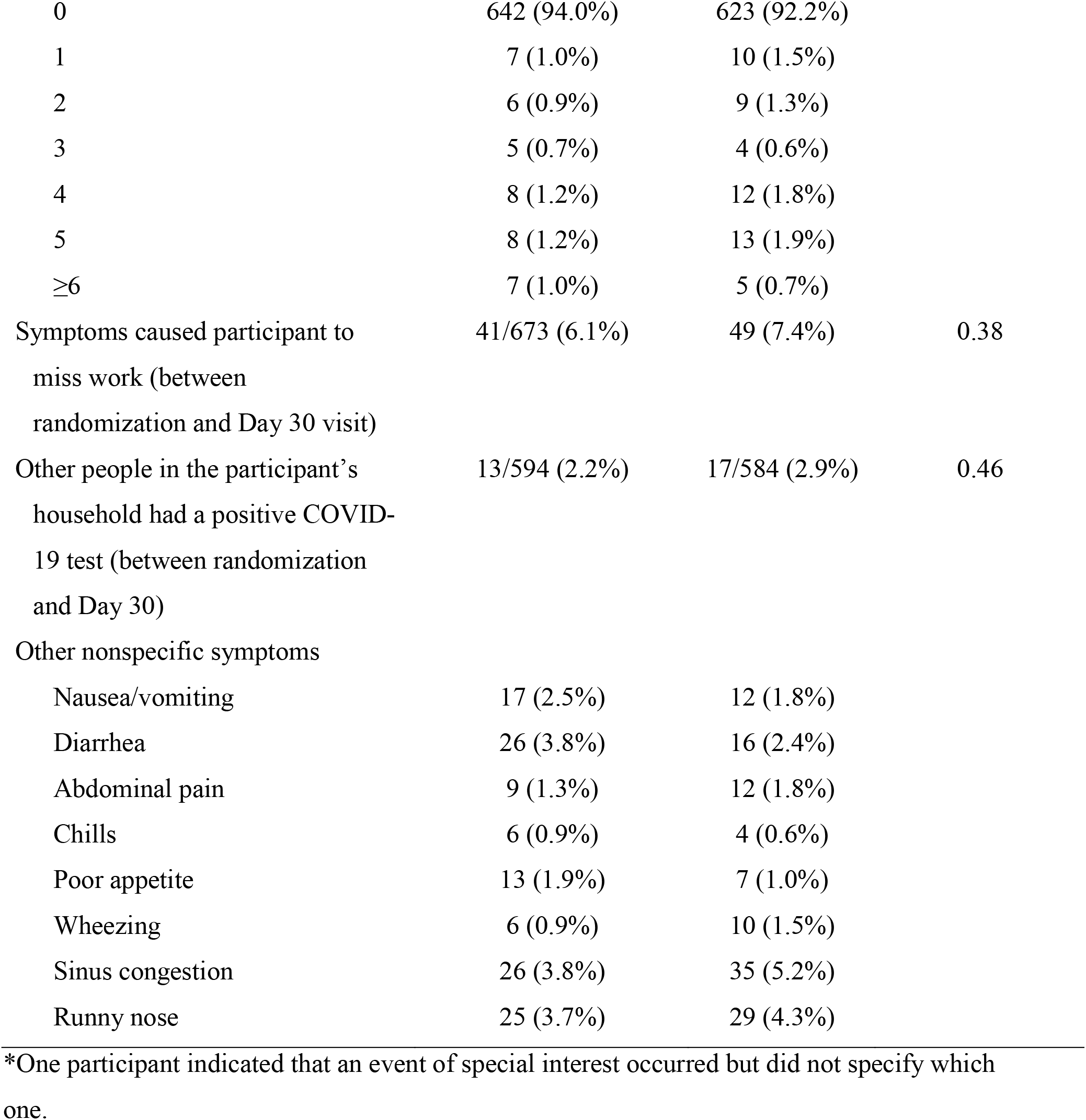
Serious adverse events, adverse events of special interest, adverse events, self-reported adherence, and self-reported symptoms.

|  | **Hydroxychloroquine**<br>**(N=683)** | **Placebo**<br>**(N=676)** | **p-value** |
| --- | --- | --- | --- |
| Number of participants with serious adverse events (to day 60) | 3 (0.4%) | 2 (0.3%) | 1.00 |
| Serious adverse event resulted in initial or prolonged hospitalization for the participant | 2 (0.3%) | 1 (0.1%) | 1.00 |
| Adverse events of special interest | 7 (1.0%) | 8 (1.2%)* | 0.80 |
| Fever | 6 (0.9%) | 1 (0.1%) |  |
| Arrhythmias (ventricular) | 1 (0.1%) | 1 (0.1%) |  |
| Psychosis | 0 | 2 (0.3%) |  |
| Angioedema | 0 | 1 (0.1%) |  |
| Prolonged QT interval | 0 | 1 (0.1%) |  |
| Secondary bacterial | 0 | 1 (0.1%) |  |
| Suicidal ideation | 1 (0.1%) | 0 |  |
| Adverse events (to Day 60) | 16 (2.3%) | 13 (1.9%) | 0.71 |
| Adherence (self-reported at 100%) | 645 (94.4%) | 647 (95.7%) | 0.32 |
| COVID-related symptoms |  |  |  |
| Fatigue | 29 (4.2%) | 35 (5.2%) |  |
| Muscle aches / joint pain | 22 (3.2%) | 26 (3.8%) |  |
| Cough | 35 (5.1%) | 46 (6.8%) |  |
| Dyspnea | 13 (1.9%) | 15 (2.2%) |  |
| Headache | 29 (4.2%) | 33 (4.9%) |  |
| Sore throat | 14 (2.0%) | 20 (3.0%) |  |
| Fever | 5 (0.7%) | 4 (0.6%) |  |
| Loss of smell | 5 (0.7%) | 4 (0.6%) |  |
| Loss of taste | 4 (0.6%) | 3 (0.4%) |  |
| Number of COVID-related symptoms |  |  |  |
| 0 | 642 (94.0%) | 623 (92.2%) |  |
| 1 | 7 (1.0%) | 10 (1.5%) |  |
| 2 | 6 (0.9%) | 9 (1.3%) |  |
| 3 | 5 (0.7%) | 4 (0.6%) |  |
| 4 | 8 (1.2%) | 12 (1.8%) |  |
| 5 | 8 (1.2%) | 13 (1.9%) |  |
| ≥6 | 7 (1.0%) | 5 (0.7%) |  |
| Symptoms caused participant to miss work (between randomization and Day 30 visit) | 41/673 (6.1%) | 49 (7.4%) | 0.38 |
| Other people in the participant's household had a positive COVID-19 test (between randomization and Day 30) | 13/594 (2.2%) | 17/584 (2.9%) | 0.46 |
| Other nonspecific symptoms |  |  |  |
| Nausea/vomiting | 17 (2.5%) | 12 (1.8%) |  |
| Diarrhea | 26 (3.8%) | 16 (2.4%) |  |
| Abdominal pain | 9 (1.3%) | 12 (1.8%) |  |
| Chills | 6 (0.9%) | 4 (0.6%) |  |
| Poor appetite | 13 (1.9%) | 7 (1.0%) |  |
| Wheezing | 6 (0.9%) | 10 (1.5%) |  |
| Sinus congestion | 26 (3.8%) | 35 (5.2%) |  |
| Runny nose | 25 (3.7%) | 29 (4.3%) |  |

\*One participant indicated that an event of special interest occurred but did not specify which one.

## DISCUSSION

### Statement of Principal Findings

The study was underpowered to meet its primary objective, and the test of the primary endpoint does not provide evidence of a benefit for HCQ for PrEP in a high-risk HCW population.

### Strengths and Weaknesses in Context

The original study design was powered to show a 20% relative treatment effect assuming a 5% event rate in the placebo arm. However, due to slowed enrollment early in the study, the study was amended to decrease the sample size and hence the power, increasing the detectable relative treatment effect to 50%. Thus, the study was underpowered to detect a small treatment effect.

Also, while the partially remote nature of the trial was novel and improved feasibility, particularly during a pandemic, it also resulted in the limitation that we did not have laboratory confirmation for COVID-19–like illness. Early in the pandemic, in some regions, testing was not performed per local policies in HCW with suspected infection and mild or moderate symptoms. These events were defined as suspected cases and were combined with the confirmed cases in the primary outcome. This also resulted in few confirmed COVID-19 infections; thus, our primary outcome was primarily suspected COVID-19 clinical infection. While the study did not have frequent PCR testing, testing at entry and end-of-study intervention showed low cross-sectional asymptomatic rates of viral shedding in the study population. Similarly, the seroconversion rate was low (0.6%) over the 30-day intervention period. These low rates of asymptomatic shedding and seroconversion suggest that SARS-CoV-2 infection was lower than expected in the study population and may be overestimated by our composite primary outcome definition.

There are strengths to highlight in the HERO-HCQ trial that might inform future clinical trials. As with many trials, there was a need for rapid development and execution; HERO-HCQ went from initial discussions with the funder to first participant enrolled in 1 month. Although there was a need to move expeditiously, HERO-HCQ did not trade speed for validity. Many studies during the pandemic have had limited impact due to significant study design limitations, particularly lack of randomization and blinding to intervention arm. HERO-HCQ is a randomized, double-blind, placebo-controlled trial. In part to respond to the need to limit in-person activities and to design a trial that was patient-centered and pragmatic, HERO-HCQ used direct-to-participant recruitment and a participant-facing portal to capture patient-reported outcomes. As recruitment and retention remain significant challenges for many clinical trials, we believe these strategies should be considered more frequently in trial design across most disease states.

Globally, there remains a role for an oral preventative for SARS-CoV-2 infection, particularly in regions where access or acceptance to preventative vaccines remains low. In April 2020, the COVID PREP team opened a randomized, double-blind, placebo-controlled clinical trial that would enroll participants from the United States and Canada to test the hypothesis that HCQ could be effective as a prophylactic treatment in the setting of HCW who have high risk of exposure to SARS-Cov2.^16^ In this completely remote, direct-to-participant trial, participants were randomized 1:1:1 to placebo or HCQ 400 mg once weekly or twice weekly by mouth for 12 weeks. Like the HERO-HCQ study, the primary endpoint was confirmed or probable COVID-19-compatible illness.

The COVID PREP study enrolled 1483 participants — 494 in the placebo group, 494 in the one-weekly dosing group, and 495 in the twice-weekly group. The pooled effect of the HCQ groups vs. placebo yielded a hazard ratio of 0.73 (95% CI 0.48 to 1.09) with a non-statistically significant p-value of 0.12. The point estimates were similar to the hazard ratio in HERO-HCQ 0.75 (95% CI 0.50 to 1.13). The HERO-HCQ and COVID PREP studies are compared in **Supplemental Table 3**. Pooling the main results using the Mantel–Haenszel method resulted in an estimate of the common odds ratio of 0.74 (95% CI 0.55 to 1.00) with a p-value of 0.046. The Breslow-Day test of homogeneity suggested that the studies were highly comparable (p-value=0.92).

Thus, pooling the studies’ data suggests potential for a modest benefit for HCQ for reducing the primary endpoint event rate by approximately 25%. Both studies had similar limitations, including the early closure that resulted in inadequate power to assess the primary endpoints and the low number of laboratory-confirmed SARS-CoV-2 infections. Both trials leveraged direct-to-participant recruitment and participant facing portals to limit in-person activities (COVID PREP was fully remote), which limited the ability to confirm COVID-19–like symptoms as SARS-CoV-2 infection. There is another difference between the two studies that requires reconciliation — the doses of HCQ used in COVID PREP and HERO-HCQ are different. COVID PREP, as noted above, used a once or twice weekly 400-mg dose, while HERO-HCQ used a daily 400-mg dose following a 600-mg twice-daily loading dose. As summarized by Rajasingham et al.,^16^ there are no validated therapeutic target concentrations for HCQ either for treatment or prevention of COVID-19, and in COVID PREP adequate plasma concentrations are not achievable to reach in vitro targets, which would be required to propose that the drug is effective for prevention of SARS-CoV-2 infection.

## Conclusion and Future Research

The prophylactic use of HCQ by HCW was safe but did not produce a clinically useful treatment to prevent COVID-19 clinical infection. These results suggest a larger randomized controlled trial or meta-analysis is needed to determine the potential for HCQ in reducing the rates of clinical infection with COVID-19.

### Summary Box

**What is already known on this topic**

- In the absence of any effective treatment or prophylactic against SARS-CoV-1, researchers noted that repurposed medications could provide protection, especially for at-risk healthcare workers (HCWs)
- In vitro data suggested hydroxychloroquine (HCQ) could have antiviral activity against SARS-CoV-1
- At the time of study initiation, no data existed on the efficacy of HCQ as a prophylactic for SAR-CoV-1, thus evidence was urgently needed

**What this study adds**

- The prophylactic use of HCQ by HCW was safe but did not produce a clinically useful treatment to prevent COVID-19 clinical infection
- The trial stopped early due to failure to enroll, thus was underpowered to answer the question about efficacy of HCQ for prophylactic benefit for SARS-CoV-1

## Supporting information

HERO Research Program Collaborators

## Data Availability

Data will be made available upon reasonable request and per the PCORI guidelines for open science and transparency.

## Acknowledgments

The authors thank the site investigators, study teams, and participants who made this project possible, and who contributed to scientific research during a time of immediate need.

## Footnotes

### Contributors

The senior author (AH) obtained funding from the Patient-Centered Outcomes Research Institute (PCORI) to conduct the study (Contract Number COVID-19-2020-001). AH and the lead author (SN) affirm the manuscript is an honest, accurate, and transparent account of the study being reported; that no important aspects of the study have been omitted, and that any discrepancies from the study as planned have been explained. SN, LC, EF, AF, RO, CW, KA, and AH conceived, designed, and oversaw the study, and led manuscript development. JG, HM, JS, and KA were responsible for data management and statistical analysis. All of the authors participated in data collection and acquisition; reviewed the manuscript for important intellectual content; and gave administrative, technical, or material support. The lead author had full access to all the data in the study and takes responsibility for the integrity, transparency of the data, and the accuracy of the data analysis. The corresponding author attests that all listed authors meet authorship criteria and that no others meeting the criteria have been omitted.

### Ethical approval

The HERO-HCQ trial was reviewed by the Duke University School of Medicine Institutional Review Board and approved by the Western Institutional Review Board (Pro00105274).

## References

1. CDC COVID-19 Response Team. Characteristics of health care personnel with COVID-19 — United States, February 12–April 9, 2020. MMWR Morb Mortal Wkly Rep 2020;69(15):477–481. doi: 10.15585/mmwr.mm6915e6.

2. Kalil AC. Treating COVID-19—off-label drug use, compassionate use, and randomized clinical trials during pandemics. JAMA 2020;323(19):1897–1898. doi: 10.1001/jama.2020.4742..

3. McCreary EK, Pogue JM. COVID-19 treatment: a review of early and emerging options. Open Forum Infect Dis. 2020;7(4):ofaa105. doi: 10.1093/ofid/ofaa105.

4. Wang M, Cao R, Zhang L, et al. Remdesivir and chloroquine effectively inhibit the recently emerged novel coronavirus (2019-nCoV) in vitro. Cell Res. 2020;30(3):269–271. doi:10.1038/s41422-020-0282-0.

5. Forrest CB, Xu H, Thomas LE, et al.; HERO Registry Research Group. Impact of the early phase of the COVID-19 pandemic on US healthcare workers: results from the HERO Registry. J Gen Intern Med. 2021;1–8. doi: 10.1007/s11606-020-06529-z.

6. Forrest CB, McTigue KM, Hernandez AF, et al. PCORnet® 2020: current state, accomplishments, and future directions. J Clin Epidemiol 2020;129:60–67.

7. Pilkonis PA, Choi SW, Reise SP, et al. Item banks for measuring emotional distress from the Patient-Reported Outcomes Measurement Information System (PROMIS®): depression, anxiety, and anger. Assessment. 2011;18(3):263–283. doi:10.1177/1073191111411667

8. West, C.P., et al., Single item measures of emotional exhaustion and depersonalization are useful for assessing burnout in medical professionals. J Gen Intern Med 2009; 24(12):1318–21.

9. Rohland BM, Kruse GR, Rohrer JE. Validation of a single-item measure of burnout against the Maslach Burnout Inventory among physicians. Stress and Health 2004;20(2):75–9. doi: 10.1002/smi.1002.

10. McMurray, J.E., et al., The work lives of women physicians results from the physician work life study. The SGIM Career Satisfaction Study Group. J Gen Intern Med 2000; 15(6): 372–80.

11. Dolan, E.D., et al., Using a single item to measure burnout in primary care staff: a psychometric evaluation. J Gen Intern Med 2015;30(5):582–7.

12. Kroenke, K., R.L. Spitzer, and J.B. Williams, The Patient Health Questionnaire-2: validity of a two-item depression screener. Med Care 2003; 41(11): 1284–92.

13. Arroll, B., et al., Validation of PHQ-2 and PHQ-9 to screen for major depression in the primary care population. Ann Fam Med 2010; 8(4): 348–53

14. Garner W. Constructing confidence intervals for the differences of binomial proportions in SAS®. Available at: https://www.lexjansen.com/wuss/2016/127_Final_Paper_PDF.pdf.

15. Cochran WG. Some methods for strengthening the common X2 tests. Biometrics. 1954;10:417–451. doi: 10.2307/3001616.

16. Rajasingham R, Bangdiwala AS, Nicol MR, et al.; COVID PREP Team. Hydroxychloroquine as pre-exposure prophylaxis for COVID-19 in healthcare workers: a randomized trial. Clin Infect Dis. 2020 Oct 17:ciaa1571. doi: 10.1093/cid/ciaa1571. Online ahead of print.

17. Rojas-Serrano J, Portillo-Vásquez AM, Thirion-Romero I, et al. Hydroxychloroquine for prophylaxis of COVID-19 in health workers: a randomized clinical trial. https://www.medrxiv.org/content/10.1101/2021.05.14.21257059v1. doi: https://doi.org/10.1101/2021.05.14.21257059

