## Supplemental File for "Hydroxychloroquine for pre-exposure prophylaxis of COVID-19 in health care workers: a randomized, multicenter, placebo-controlled trial (HERO-HCQ)"

**Supplemental Figure 1.** HERO Registry and HERO-HCQ Trial Timeline

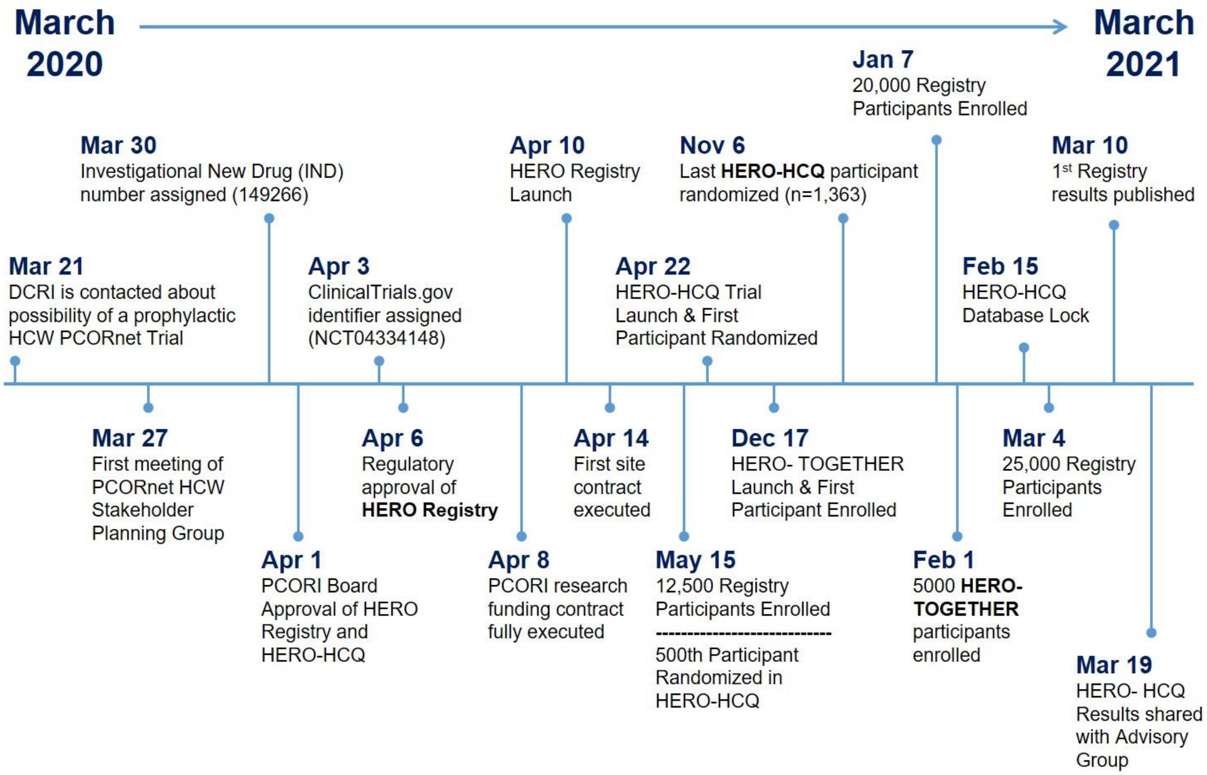

**Supplemental Table 1.** Participating PCORnet HERO-HCQ Trial Sites

| |
|---|
| AdventHealth Orlando |
| Allina Health |
| Baylor Scott and White Memorial Hospital |
| Children's Hospital Colorado |
| Clinical Trials Center of Middle Tennessee |
| Columbia University Medical Center |
| Duke University Hospital |
| Hospital for Special Surgery |
| Johns Hopkins Hospital |
| Marshfield Clinic |
| Mayo Clinic |
| Michigan Medicine |
| New York Presbyterian/Weill Cornell Medical Center |
| Northwestern University |
| Ochsner Clinic Foundation |
| Rush University Medical Center |
| Seattle Children's Hospital |
| Tampa General Hospital |
| Temple University Hospital |
| The Ohio State University Wexner Medical Center |
| University Medical Center- New Orleans |
| University of Florida Health |
| University of Florida Health Central Florida |
| University of Florida Jacksonville Shands Medical Center |
| University of Iowa |
| University of Kansas Medical Center |
| University of Miami Miller School of Medicine |
| University of Missouri - Columbia |
| University of Nebraska Medical Center |
| University of North Carolina at Chapel Hill |
| University of Pittsburgh School of Medicine |
| University of Texas Southwestern Medical Center Dallas |
| Vanderbilt University Medical Center |
| Wake Forest Baptist Medical Center |

**Supplemental Table 2.** Comparison of Confidence Interval (CI) Estimates for the Primary Analysis

| **CI Method** | **95% CI (% scale)** |
|---|---|
| Miettinen-Nurminen | -4.60, 0.87 |
| Chang-Zhang | -4.61, 0.90 |
| Agresti-Caffo | -4.55, 0.88 |
| Hauck-Anderson | -4.61, 0.94 |
| Miettinen-Nurminem-Mee | -4.60, 0.87 |
| Newcombe | -4.59, 0.88 |
| Newcombe (corrected) | -4.69, 0.99 |
| Wald (corrected) | -4.68, 1.01 |

Note: the point estimate is -1.84% favoring HCQ vs. placebo at Day 30. The 2-sided Fisher's exact p-value is 0.20.

**Supplemental Table 3.** Comparison of COVID PREP and HERO-HCQ studies

| | **COVID PREP (NCT04328467)** | **HERO-HCQ (NCT04334148)** |
|---|---|---|
| Randomization ratio | 2:2:1:1<br>(the placebo arms were pooled) | 1:1 |
| HCQ dosage | Loading dose of 400 mg (two 200-mg tablets) twice separated by 6-8 hours followed by (i) 400 mg (two 200-mg tablets) **once** weekly for 12 weeks or (ii) 400 mg (two 200-mg tablets) **twice** weekly for 12 weeks | 600-mg loading dose of study drug twice on the first day, followed by 400 mg daily for 29 days |
| Maximum follow-up | 3 months | Treatment period for 30 days with follow-up at Day 60 |
| Planned sample size | ~3,150 HCW<br>(1050 per arm) | 15,000 HCW<br>(7500 per arm) |
| Final sample size<br>(% of planned) | 1483<br>(47.2%) | 1359<br>(9.1%) |
| HCQ primary endpoint event rate | 58/989 (5.9%) | 41/683 (6.0%) |
| Placebo primary endpoint event rate | 39/494 (7.9%) | 53/676 (7.8%) |

Mantel–Haenszel Common Odds Ratio = **0.74 (95% CI 0.55 to 1.00), p= 0.046.** Breslow-Day Test of Homogeneity p-value= 0.92.
