## Supplementary material for "Hydroxychloroquine for pre-exposure prophylaxis of COVID-19 in health care workers: a randomized, multicenter, placebo-controlled trial (HERO-HCQ)": HERO Research Program Collaborators

| | | |
|---|---|---|
| Rachel | Addison | |
| Amanda | Adler | |
| Faraz | Ahmad | |
| Hailey | Anderson | |
| Brandon | Apagueno | |
| Khaula | Baloch | |
| Adrienne | Baranauskas | |
| Allison | Beaver | |
| Cassidy | Bowen | |
| Jennifer | Bowman | |
| Ross | Boyce | |
| Abigail | Boyer | |
| Emily | Bozant | |
| Amber | Brock | |
| Julie | Castex | |
| Matthew | Catania | |
| Lyndsay | Christensen | |
| Caitlin | Clohessey | |
| Jennifer | Cook | |
| Kirsty | Cowles | |
| Judith | Currier | |
| Vanessa | Curtis | |
| Tami | Day | |
| Mihiri | De Silva-Udawatta | udawatta |
| Vladimir | Demyanenko | |
| Brenda | Farlow | |
| Kristen | Foss | |
| Amy | Franklin | |
| Ryan | Fraser | |
| Sandra | Freeman | |
| Jennifer | Frey | |
| Mary | Froilan | |
| Jyotsna | Fuloria | |
| Varsha | Gajjar | |
| Sarah | Galloway | |
| Dianne | Gallup | |
| Carol | Geary | |
| Eden | Gebre | |
| Nina | Gentile | |

| | | |
|---|---|---|
| Alison | Gimbel | |
| Leigh | Gosnell | |
| Brett | Gray | |
| Tina | Harding | |
| Robert | Haws | |
| Loreen | Herwaldt | |
| Peter | Higgins | |
| Kathy | Hijek | |
| Zhen | Huang | |
| Jenny | Jackman | |
| Dushyantha | Jayaweera | |
| Patricia | Karausky | |
| Stephanie | Katz | |
| Brenda | Lane | |
| Jeffrey | Leimberger | |
| Renee | Leverty | |
| Kelly | Lindblom | |
| Brittney | Manning | |
| Mike | Mantell | |
| Nizar | Maraqa | |
| Jennifer | Martin | |
| M. Patricia | McAdams | |
| James | McClay | |
| Brian | McCourt | |
| Steve | McNulty | |
| Jenna | McPhee | |
| Lisa | Merck | |
| Maggie | Messplay | |
| Brenda | Mickley | |
| Bethany | Miller | |
| Karen | Miller | |
| Caryn | Morse | |
| M Hassan | Murad | |
| Syed | Naqvi | |
| Celia | Nelson | |
| Matthew | Newell | |
| Jenn | Parrish | |
| Trish | Perl | |
| Thomas | Philips | |
| Joanna | Pomerantz | |

| | | |
|---|---|---|
| Kelli | Porzondek | |
| Vidya | Raghavan | |
| Sarah | Ramey | |
| Suchitra | Rao | |
| Hannah | Reimer | |
| Frank | Rhame | |
| Yoona | Rhee | |
| Patricia | Robinson | |
| Frank | Rockhold | |
| Lisa | Rohn | |
| David | Sielaty | sielaty |
| Jeremy | Smith | |
| Phillip | Smith | |
| Lauren | Southerland | |
| Leanne | Stanton | |
| Becky | Staub | |
| Emily | Stein | |
| Theresa | Strakos | |
| Mark | Sulkowski | |
| Martha | Summerlin | |
| Thomas | Tanner | |
| Stephanie | Taylor | |
| Deborah | Theodore | |
| James | Topping | |
| Jessica | Wallan | wallan |
| Mark | Ward | |
| Laura | Webb | |
| Jun | Wen | |
| Amy | West | |
| Jo | Wheeler | |
| Timothy | Wilkin | wilkin |
| Adeia | Williams | |
| Amelia | Williams | |
| Quintara | Williams | williams |
| Anne | Wolfley | |
| Weibing | Xing | |
| Qinghong | Yang | |
| Pearl | Zakroysky | |
| Caroline | Zaworski | zaworski |
| Danielle | Zerr | zerr |

| | | |
|---|---|---|
| Songlin | Zhu | |
| Elizabeth | Zieser | misenheimer |
